# Exome-wide association study reveals 7 functional variants associated with ex-vivo drug response in acute myeloid leukemia patients

**DOI:** 10.1101/2023.08.02.23290523

**Authors:** Anil K Giri, Jake Lin, Konstantinos Kyriakidis, Garima Tripathi, Henrikki Almusa

## Abstract

Acute myeloid leukemia (AML) is a rare aggressive blood cancer without any long-term cure. Further, due to the extreme molecular heterogeneity of the disease, drug treatment response varies from patient to patient. The variability of drug response can cause unnecessary treatment in more than half of the patients with no or partial therapy responses leading to severe side effects, economic as well as time loss. Understanding the genetic risk factors underlying the drug response in AML can help with improved prediction of treatment responses and identification of biomarkers in addition to mechanistic insights to monitor treatment response.

Here, we report the results of the largest exome-wide association study (EWAS) of ex-vivo drug response performed to date with 175 AML cases and 47 drugs. We used information from 55423 exonic SNPs to perform the analysis. We identified exome-wide significant (p<9.02 ×10^-7^) associations for rs113985677 in *CCIN* with tamoxifen response, rs115400838 in *TRMT5* with idelalisib response, rs11878277 in *HDGFL2* with entinostat, and rs2229092 in LTA associated with vorinostat response.

Further, using multivariate genome-wide association analysis, we identified the association of rs11556165 in *ATRAID*, and rs11236938 in *TSKU* with the combined response of all 47 drugs and 29 nonchemotherapy drugs at the genome-wide significance level (p<5×10^-8^). Additionally, a significant association of rs35704242 in *NIBAN1* was associated with the combined response for nonchemotherapy medicines (p=2.51×10^-8^), and BI.2536, gefitinib, and belinostat were identified as the central traits. Our study represents the largest EWAS study to date on ex-vivo drug response in AML and reports 7 new associated loci that help to understand the anticancer drug response in AML patients.

## Introduction

Acute myeloid leukemia (AML) is a biologically heterogeneous aggressive hematological malignancy^1,2^. The inherent heterogeneity not only influences the clinical presentation of the disease but also affects the response and toxicity towards a therapy^3,4^. Hence, huge interindividual variability exists for drug response in AML patients^5,6,7^.

The variability in the treatment response of AML patients is driven by a broad spectrum of genetic (e.g., somatic and germline variants) and nongenetic alterations (e.g., age) present in different patient groups.^8^ Although the somatic mutations affecting treatment response in AML have been extensively studied in AML^9,10^, the germline variants contributing to the drug response largely remain under-explored mostly because of a lack of well-characterized cohorts with both genetic and drug treatment data^11-13^. Further, the number of drug-treatment that can be explored in a clinical cohort is limited because patients are usually treated with one or two common drugs approved in the clinics, hence neglecting the other drugs. Furthermore, the collection of clinical responses to drugs in a large patient cohort will require long follow-up time, resources, and a network that could be beyond the capability of most academic laboratories.

The drug sensitivity and resistance testing (DSRT) platform can measure the ex-vivo response for hundreds of drugs at multiple doses in patient-derived primary cells^14-16^. The ex-vivo drug response can provide a reasonable estimate for the clinical response of the patient towards the drug^3,9^ and has been increasingly used in guiding the treatment for refractory/relapse patients with no available standardized treatment. Further, the ex-vivo drug response measured in patient-derived-primary samples has been used as a surrogate for clinical response to identify genetic variants affecting treatment response in a patient.

Here, using the germline information from the exome-sequence and ex-vivo response data from 47 drugs measured in 75 AML patients, we performed an exome-wide association study to identify functional variants affecting drug response in AML. The result identifies 4 novel loci near *CCIN*, *TRMT5*, *HDGFL2*, and *LTA* genes affecting the ex-vivo response of tamoxifen, idelalisib, entinostat, and vorinostat in AML at exome-wide significance levels. Further, we also used a novel and efficient statistical framework “MetaPhat” to conduct and decompose multivariate GWAS signals affecting ex-vivo drug response in AML patients. The analysis identified the association of loci in *NIBAN1/*, *ATRAID*, and *TSKU* with combined drug response in AML.

## Methods

### AML Patient’s cohort

One seventy-five bone marrow aspirates and peripheral blood samples (leukemic cells) and skin biopsies (noncancerous cells for germline genomic information) from high-risk AML patients were obtained after informed consent with approval (No. 239/13/03/00/2010, 303/13/03/01/2011). Patient characteristics are summarized in Supplementary Table 1.

### Exome-sequencing of the samples

Genomic DNA was isolated from the skin biopsies samples using the DNeasy Blood & Tissue Kit (Qiagen) and was used for exome sequencing. For exome sequencing, 3 μg DNA was fragmented and processed according to the NEBNext DNA Sample Prep Master Mix protocol. Exome capture was performed using the Nimblegen SeqCap EZ v2 capture Kit (Roche NimbleGen). Sequencing of exomes was done using HiSeq1500, 2000, or 2500 instruments (Illumina), and approximately 4 × 10^7^ bp paired-end reads were sequenced per sample. The data was curated and analyzed as described before^9, 17^.

Briefly, the sequenced raw exome reads were trimmed using Trimmomatic (version 0.36) to remove adapters and low-quality segments. The trimmed data were aligned to the human reference genome (GRCh build 37) using Burrows-Wheeler Aligner (bwa version 0.7.12, Wellcome Trust Sanger Institute, Cambridge, UK). After alignment, the potential PCR duplicates were removed using Picard (Broad Institute of Harvard and MIT, Cambridge, MA, USA), and BAM files were sorted, and indexed using SAMtools (Wellcome Trust Sanger Institute, Cambridge, UK). From the aligned reads, GATK (version 4.1.3.0) was used to joint call the samples for genetic variants. Variant annotation was performed using ANNOVAR and Ensembl version 6851.

We performed a stringent quality control analysis of the genetic data (Supplementary Figure 1). We removed 1 sample with a genomic call rate of less than 95%, and 9 samples with missing information for >10 of the drug response. We also removed variants without proper annotation (without rsid), call rate (<90%), MAF <0.01, and Hardy Weinberg equilibrium p-valueC<C10^−5^) were removed.

#### Drug resistance and sensitivity testing

We measured ex-vivo response for a library of 47 commercially available chemotherapeutic and targeted oncology compounds consisting of 34 approved drugs, and 13 investigation compounds (Supplementary Table 2) using the DSRT platform at the high throughput biomedicine unit at FIMM as described before^14,18^. Briefly, drugs were plated in a 10-fold dilution series of five concentrations covering a 1,000-fold concentration range on clear-bottomed 384-well plates (Corning #3712), using an Echo 550 Liquid Handler (Labcyte). The concentration ranges were selected for each compound separately to investigate their full dynamic range of the dose-response relationships.

Next, 20 μl of AML cells (approximately10,000) was added to a pre-drugged plate containing the mononuclear cell medium (MCM; PromoCell) supplemented with 0.5 μg/mL gentamicin and 2.5 μg/mL amphotericin B^3,18^. The cell was maintained in the culture media at 37 °C with 5% CO2 in a humidified incubator. After 72h incubation, cell viability was measured using CellTiter-Glo (Promega) reagent in duplicate using a PHERAstar FS (BMG Labtech) plate reader. As positive (total killing) and negative (non-effective) controls, we used 100μM benzethonium chloride and 0.1% dimethyl sulfoxide (DMSO), respectively, when calculating the relative inhibition percent.

The percentage inhibition was calculated by normalizing the cell viability to negative control wells containing only 0.1% dimethyl sulfoxide (DMSO) and positive control wells containing 100 μM cell-killing benzethonium chloride (BzCl). The drug responses passing the data quality assessment were included in further analysis. Drug sensitivity score (DSS), a measure of drug efficacy for each drug was calculated for each patient as described before^19^. Next, selective DSS (sDSS) was calculated by normalizing drug responses against 17 healthy controls.

#### Single SNP exome-wide association analysis

We only selected 47 drugs for an analysis whose ex-vivo response was measured in all the patients. The sDSS scores for each drug were inverse normalized using the inbuilt command in R (http://www.r-project.org/). We performed a linear regression model using PLINK^20^ to test the association of SNPs with inverse normalized sDSS. Cell culture medium for the drug assay, disease stage (relapse/diagnosis), and first five principal components were used as covariates in the model. All other statistical analyses and plotting were performed using R programming. We used the Bonferroni correction for multiple tests (n = 55423 tests) and set the statistical significance and suggestive threshold to P-values less than 9.02 ×10^−7^ and 1×10^−4^, respectively.

#### Multivariate exome-wide association analysis using MetaPhat

Several of the drugs measured in the experiment have a similar mechanism of action and act through the same pathway. As these functionally similar drugs have a correlated response in the studied samples, we used our novel method to conduct multivariate EWAS of a linear combination of correlated traits using univariate GWAS for 55423 SNPs results as implemented in MetaPhat^21^ (integrating metaCCA^22^). Further, we decomposed the multivariate associations into sets of central biomarker traits to understand the possible mechanism of action for the identified variant. This analysis tested each SNP separately for its simultaneous associations with all 47 phenotypes, 18 chemotherapies, and 29 nonchemotherapies drugs. A multivariate approach had the dual advantages of achieving data reduction and increasing statistical power compared with running 47 separate univariates EWAS.

Since we did not have a replication cohort, we performed a statistical experiment to validate the true nature of identified association in multivariate EWAS by estimating the chance of observing such an association in a random dataset. For this, we assigned random patient identity to each row of the phenotype file while keeping the correlation between the drug response the same and performed the univariate and multivariate association analysis with the genotypes using the same parameters.

## Results

### Single SNP association with the ex-vivo drug response identified association of 4 locus at exome-wide significance level

After stringent quality control, we analyzed a total of 55423 genetic markers for their association with single-drug response in 175 AML patients using linear regression adjusting for gender, age (at diagnosis for cases), cell culture medium, and the first 10 principal components. We detected an exome-wide significance level association for rs113985677 in *CCIN* with Tamoxifen (p=3.68×10- 07), and rs115400838 in *TRMT5* with the idelalisib response (p=4.20×10^-7^). We also observed an EWAS level significance for rs11878277 in *HDGFL2* with entinostat (p=4.94×10^-7^), and rs2229092 in *LTA* with vorinostat response (p=8.28×10^-7^) (Table 1). The distribution of the association p-value across different chromosomes have been shown in Figure 1.

**Figure 1:**
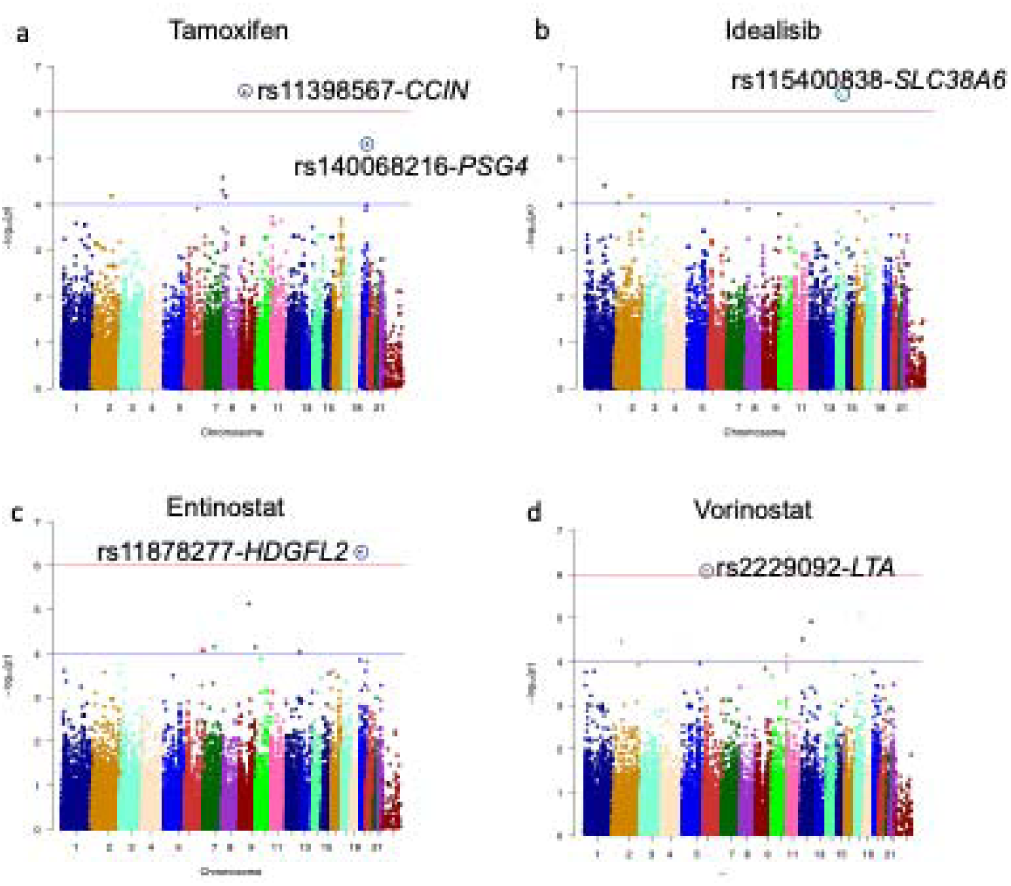
Manhattan plot depicting the -log(10) p-values for SNPs associated with (a) tamoxifen (b) idelalisib, (c) entinostat, and (d) vorinostat across different chromosomes. *The p-value has been obtained using linear regression adjusted for disease stage (diagnosis/relapse), cell culture medium and first 5 principal components from genotype data*.

**Table 1:** SNPs showing association with ex-vivo drug response at exome-wide significance levels in the analysis.

| CHR | POS | SNP | Position | Gene | A1/A2 | N | Effect size | P | Phenotype | Mechanism | Approval status |
| --- | --- | --- | --- | --- | --- | --- | --- | --- | --- | --- | --- |
| 9 | 36170290 | rs113985677 | Missense<br>R (Arg) ><br>Q/p<br>(Gln/Pro) | *CCIN* | A/G | 175 | 4.45 (0.84) | $3.68 \times 10^{-7}$ | Tamoxifen | Selective<br>estrogen<br>receptor<br>modulator | Approved |
| 14 | 61446147 | rs115400838 | Missense<br>R (Ser) > P<br>(Cys) | *TRMT5* | A/T | 175 | 7.58(1.44) | $4.20 \times 10^{-7}$ | Idelalisib | PI3K<br>inhibitor | Approved |
| 19 | 4498000 | rs11878277 | Synonymous<br>S (Ser) > S<br>(Ser) | *HDGFL2* | A/G | 175 | 10.23(1.96) | $4.94 \times 10^{-7}$ | Entinostat | HDAC<br>inhibitor | Phase 2 |
| 6 | 31572980 | rs2229092 | Missense<br>H (His) > P<br>(Pro) | LTA | C/A | 173 | -4.9(1) | 8.28x10<sup>-7</sup> | Vorinostat | HDAC<br>inhibitor | Approved |

*Chromosomal positions of SNPs are based on National Center for Biotechnology Information genome build 38. The effect size was calculated with respect to the minor alleles. The association results presented were obtained from genotyped data in 175 patients.*

A good agreement was observed between the theoretical p-value distribution and calculated p-values for these drugs as shown in Supplementary Figure 2. The genomic inflation factor (λ) for the fitted model ranges between 0.98-1.01 which indicates the homogeneity of analyzed samples.

### Suggestive association in exome-wide association analysis

Further, we observed associations of SNPs with the response of multiple drugs with a similar mechanism of action at the sub-EWAS level of significance (P<10^-4^) suggesting a true nature of the association. For example, we observed the association of SNPs in the same genes with drugs having similar mechanisms of action. For example, rs2394516 and rs2394516 in *OR2J1* (olfactory receptor 2J1) were associated with both cladribine (purine analog) and clofarabine (another purine analog) suggesting a similar mechanism for influencing the drug response by identified variants (Supplementary Table 3).

Similarly, associations of rs201274224 in *PIK3CD* were observed with daunorubicin (topoisomerase II inhibitor), idarubicin (topoisomerase II inhibitor), and fludarabine (a purine analog) response suggesting a common mechanism of action probably through DNA damage and repair pathways. In total, we observed 295 suggestive associations within 248 unique SNP associated with the 47-drug response (Supplementary Figure 3, Supplementary Table 3).

### Multi-variate association analysis identified an association of 4 additional SNPS at the exome-wide significance level

In the multivariate analysis, first, we checked the association of 55423 SNPs with a combined response of 47 drugs (Supplementary Figure 4) and observed a GWAS level association for rs11236938-G/A (*TSKU*) with the combined response (p=2.51×10^-9^, Figure 2a, Supplementary Table 4). Further, the decomposition of multivariate association identified tamoxifen, BI.2536, and belinostat as the central traits, as defined by optimal Bayesian Information Criteria (BIC)^23^ subset and union with traits iteratively driving significant p-value for the multivariate association. Further, the analysis also identified an association of rs11556165-*ATRAID* (p=4.26×10^-8,^ Supplementary Table 4) with the combined response of belinostat, fingolimod, and alvocidib as the central traits at the genome-wide significance level.

**Figure 2:**
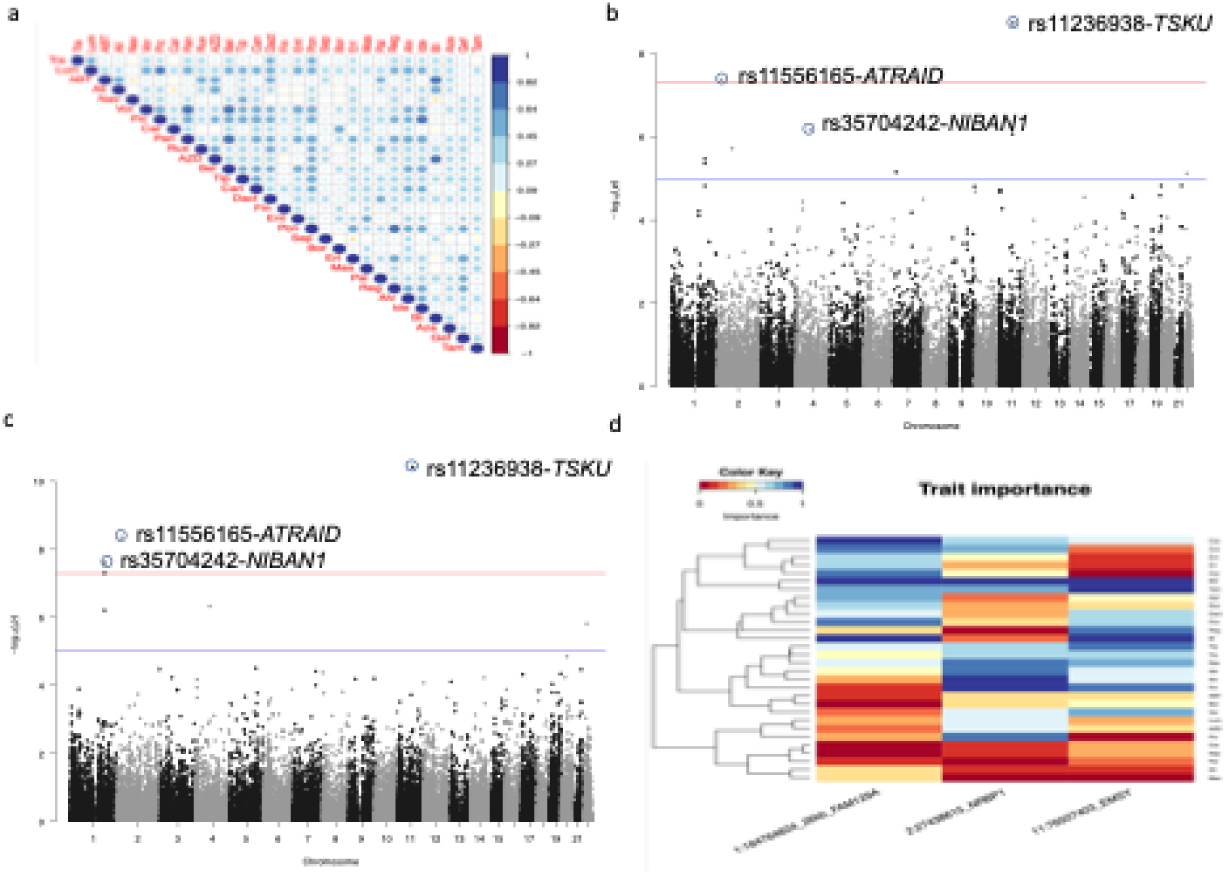
Multivariate association analysis of exonic SNPs with the linear combination of drug response. (a) Heatmap showing the correlation between the analyzed drugs based on the response from the 175 patients. (b) Manhattan plot depicting the -log (10) p-values for SNPs associated with all 47 drugs across different chromosomes in multivariate analysis**. (c)** Manhattan plot depicting the -log (10) p-values for SNPs associated with only kinase inhibitors (d)*Heatmap showing the importance of each kinase inhibitor response on the association of identified signals (The increase of importance has been shown from red to blue)*.

Next, we also performed multivariate analysis to check the association of SNPs with only a group of drugs with specific mechanisms of action (e.g., kinase inhibitors). The analysis of response from the 29 non-chemotherapy drugs identified an association of rs11236938-G/A in *TSKU* with combined drug response and tamoxifen, BI.2536, and belinostat (p=3.8×10^-11^, Figure 2b, Table 2) as the central trait. Further, rs11556165-*ATRAID* was also associated with EWAS significance level (p=3.98×10^-9^) with the combined response of non-chemo drugs with belinostat, fingolimod, alvocidib ass the major trait. Similarly, rs35704242-C/T in *NIBAN1* was associated with the combined nonchemotherapy drug response and, BI.2536, gefitinib, and belinostat as the main traits (p=2.51×10^-8^). The observed inflation factor for the p-values were 1.3 for all drug analysis and 1.17 for only nonchemotherapy drug response analysis (Supplementary Figure 5)

**Table 2:**
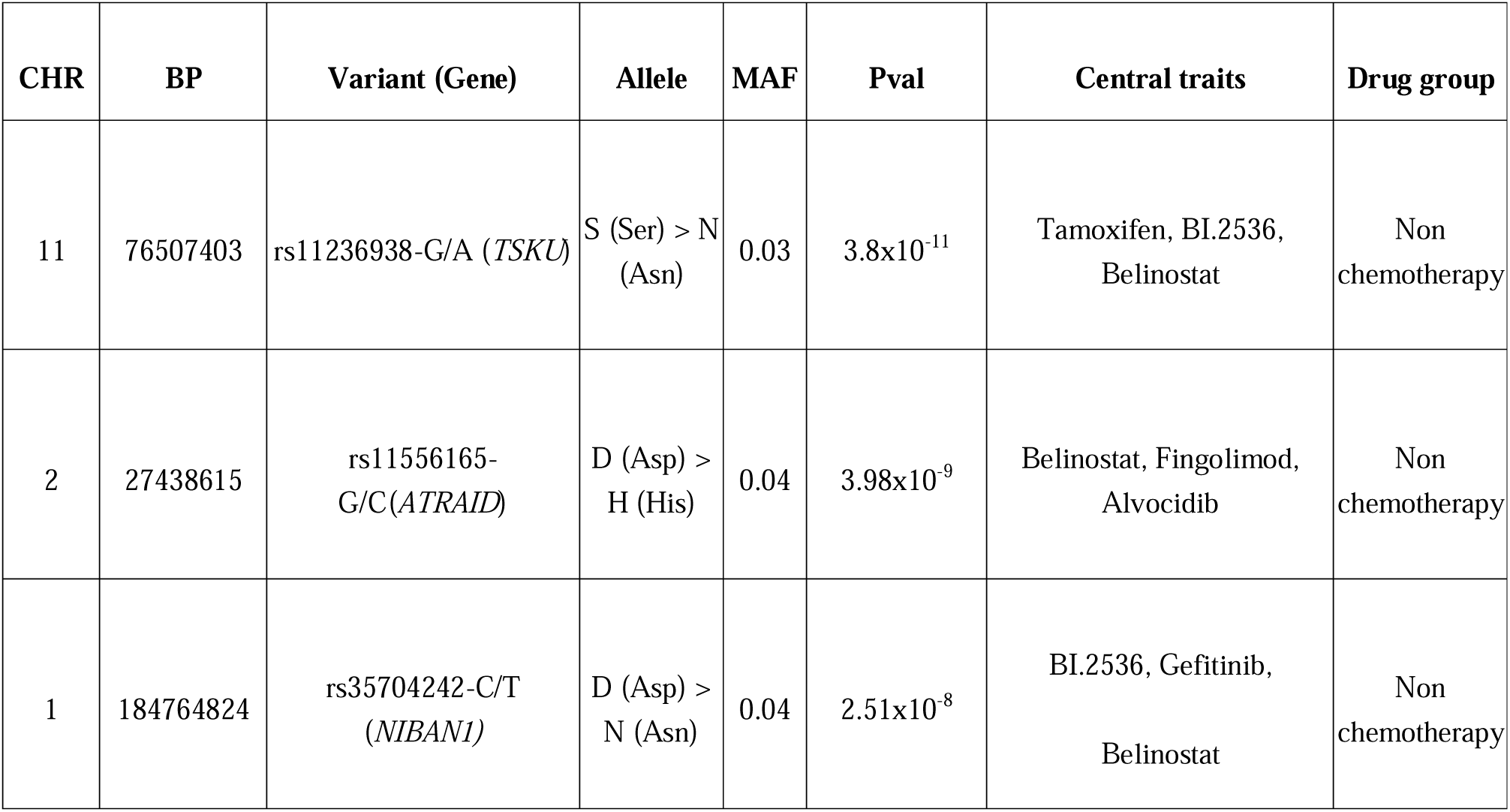
SNPs showing association with ex-vivo response of nonchemotherapy drugs as group at geome-wide significance levels in the multivariate analysis.

#### Validation of the identified association

To check the validity of the association and significance level of the p-value, we also performed an association analysis by creating 200 sets of randomized phenotypes (200×47 phenotypes) where we kept the correlation between responses of the 47 drugs the same. The univariate association analysis did not identify any signals at the exome-wide significance level at any of the random phenotypes.

However, only 8 out of 200 times any exome-wide significant association was observed when the response of all 47 drugs was analysed in the multivariate analysis. Similarly, only 5 EWAS-level significant associations were observed out of 200 times when only the response of 29 non-chemo drugs were analysed together. These analyses firmly ascertain the true nature of identified associations and revealed a maximum of 4% chance of the identified signals as false positive.

## Discussion

Genetic risk factors affecting the response of most cancer drugs are poorly defined^24,25^. Most of the large-scale genomics study in AML has explored the effect of somatic mutations on drug response undermining the influence of germline variant except for a few drugs^26,27^. Further, these studies exploring the effect of the germ-line variants on drug response have only analyzed the polymorphisms in genes encoding transporters, metabolizers, or molecular targets of chemotherapy agents neglecting most parts of the genomes^12,28^. Although specific somatic mutations have been associated with the drug response for some of the targeted therapy, such mutations account for only a minority of cases and the great majority of patients does not have such clear marker^27^. The large amount of time and resources required to collect the clinical response data has been one of the major obstacles to performing large, genome-wide association studies to identify pharmacogenetic variants affecting drug response. Our study, which comprised over 175 AML cases profiled for an ex-vivo response of 47 drugs, represents one of the largest such studies performed to date.

We identified 4 EWAS significant variants affecting the ex-vivo drug response in our cohort overall using the univariate association analysis. However, we could not validate these signals because of the lack of other cohorts having this kind of data. We identified the association of rs113985677, a missense variant (changes arginine to glutamine or proline) in *CCIN* (calcin) with Tamoxifen ex-vivo response. Calcin is a cytoskeleton protein and has been associated with lateral sinus thrombosis, spermatogenic Failure 9, globozoospermia^29,30^ and is required for the formation of the sperm head and maintain the nuclear structure in mice^31^. Downregulation of the cytoskeleton genes, connexin 43, induces tamoxifen-resistance in breast cancer^32^ cells via activation of c-Src/PI3K/Akt signaling and Epithelial to mesenchymal transition (EMT) of cells. The identified variant in the calcin gene may alter the tamoxifen response by altering the cytoskeleton dynamics during EMT.

Further, the variant is an expression quantitative trait locus for *GLIPR2* gene in the esophagus and salivary gland (P=6.5×10^-12^, and 7.9×10^-8^) where the presence of alternate allele is associated with increased expression of *GLIPR2*. *GLIPR2* is particularly expressed in leukocytes and has a role in EMT transition^33^, and autophagy^34^, suggesting a possible role in drug response via these mechanisms. However, a functional study will be needed to understand the exact role of genes and variants.

We also found a significant association between rs115400838, an upstream variant in *SLC38A6*, and a missense variant in *TRMT5* with an idelalisib response. *SLC38A6* is a glutamine and glutamate transporter^35^ and has been associated with angiotensin-converting enzyme inhibitor-induced cough^36^. The available evidence suggests that SLC38A6 may be a direct transporter of idelalisib itself or its substrates. We also observed a significant association of rs11878277 (*HDGFL2*) with entinostat in the univariate analysis. Entinostat is a histone deacetylase inhibitor and is under clinical trial for multiple cancers. *HDGFL2* (Hepatoma-Derived Growth Factor-Related Protein 2) is dispensable for Mixed Lineage Leukemia (MLL)-rearranged (MLL-R) leukemogenesis^37^ and regulates gene expression via HRP2-DPF3a-BAF complex formation^38^.

Similarly, a significant association was observed for a missense variant rs2229092 (H (His) > P (Pro) in Lymphotoxin Alpha (*LTA*) gene with vorinostat response, an HDAC inhibitor. LTA, formerly known as tumor necrosis factor-beta (TNF-β) is a toxin produced from lymphocytes (e.g., T and B) that can kill other cell types. It acts as a causative factor for autocrine and paracrine activation of canonical and noncanonical NF-κB signaling as well as promotes JAK2/STAT6 signaling in Hodgkin lymphoma^39^ resulting in prevention of apoptosis. Furthermore, LTA shapes the expression of lymphoid and myeloid-specific genes in blood. Given the wide role of *LTA* in immunity and tumor survival^40^, LTA may influence vorinostat response by modulating cancer survival-related gene expression in synergy with vorinostat.

The multivariate analysis identified rs11556165-*ATRAID* as one of the significant SNPs at EWAS level. ATRAID (all-trans retinoic acid-induced differentiation factor), is a genetic target of nitrogen-containing-bisphosphonates (N-BP), a commonly used drug to treat osteoporosis and other bone-related diseases^41^,^33^. Further, ATRAID binds with SLC37A3 (solute carrier family 37 member A3) which is a lysosomal membrane protein with 12 transmembrane segments and helps in NBP transport.

The multivariate analysis identified rs11556165-*CAD* (carbamoyl-phosphate synthetase 2, aspartate transcarbamylase, and dihydroorotase) as one of the significant SNPs at EWAS level (Table 2). CAD protein is a multifunctional multi-domain enzyme involved in the initial three speed-limiting steps of pyrimidine nucleotide synthesis. CAD is hyperactivated in most of the cancer and targeting it inhibits cancer cell survival, and self-renewal^41^.

Genetic target of nitrogen-containing-bisphosphonates (N-BP), a commonly used drug to treat osteoporosis and other bone-related diseases^41,42^. Further, ATRAID binds with *SLC37A3* (solute carrier family 37 member A3) which is a lysosomal membrane protein with 12 transmembrane segments and helps in NBP transport^41^.

We also identified the association of rs35704242 in niban apoptosis regulator 1 (NIBAN1) multi-drug response with BI 2536, gefitinib, and belinostat as the central trait set. *NIBAN1* protein is highly expressed in multiple cancers such as such as renal, colorectal, and thyroid cancer^43,44^ and may play an important role in cell survival during cellular stress such as ultraviolet irradiation cell migration and proliferation^43^, cell autophagy^45^ and immune microenvironment^46^. The association of BI 2536, gefitinib, and belinostat central responses which induce induces cell cycle, and apoptosis of cancer cells with *NIBAN1* align with the versatile function of NIBAN1 in cellular homeostasis seems justifiable.

We also observed a significant association of tsukushi (*TSKU*), a small leucine-rich proteoglycan, with overall kinase (p=2.51 x10-8) with Tamoxifen, BI.2536, Belinostat as the central traits. Tsukushin is an extracellular matrix protein and is also a hepatokine. TSKU is involved in different signaling pathways including BMP pathway, notch pathway, transforming growth factor-β (TGF-β) pathway, mitogen-activated protein kinase (MAPK)/fibroblast growth factor (FGF) pathway, and β-catenin/Wnt pathway^47^. Absence of TSKU increases cleaved poly ADP-ribose polymerase (PARP) protein levels and caspase 3/7 activity thereby increasing carcinogenicity in neuroblastoma cells^48^. Further, *TSKU* expression is strongly associated with poor overall survival in lung cancer^49^. Given its role in carcinogenicity and tumor survival, TSKU may affect drug response in cancer. However, further functional study is needed to reveal the exact mechanism by which TSKU affects drug response in AML.

In summary, our exome-wide analysis identified a potential genetic locus affecting the ex-vivo drug response of some of the common cancer therapy. These loci are in proximity to genes with a possible role in the transport of the respective drugs inside the cell. Our results provide a basis for initial exploration of the role of genes associated with this locus. However, replication studies to confirm the observed associations are needed in larger patient-group. However, the lack of another cohort for validation is a major limitation of the study. Though being a novel and significantly large cohort for a rare disease like AML, our study is still smaller than genome-wide association studies reported in other malignancies and is potentially limited in its power to detect associations. Additionally, in light of the increasing evidence that AML tumors are genetically heterogeneous, genetic predisposing factors likely vary across patients. Although we attempted to address this potential weakness by performing correlation based MetaPhat analysis, these analyses are based on drugs with different targets and mechanisms of action, resulting in the potential to miss associations due to the lack of statistical power.

## Data Availability

All data produced in the present study are available upon reasonable request to the authors

## SupplementaryInformation

**Supplementary Figure 1:**
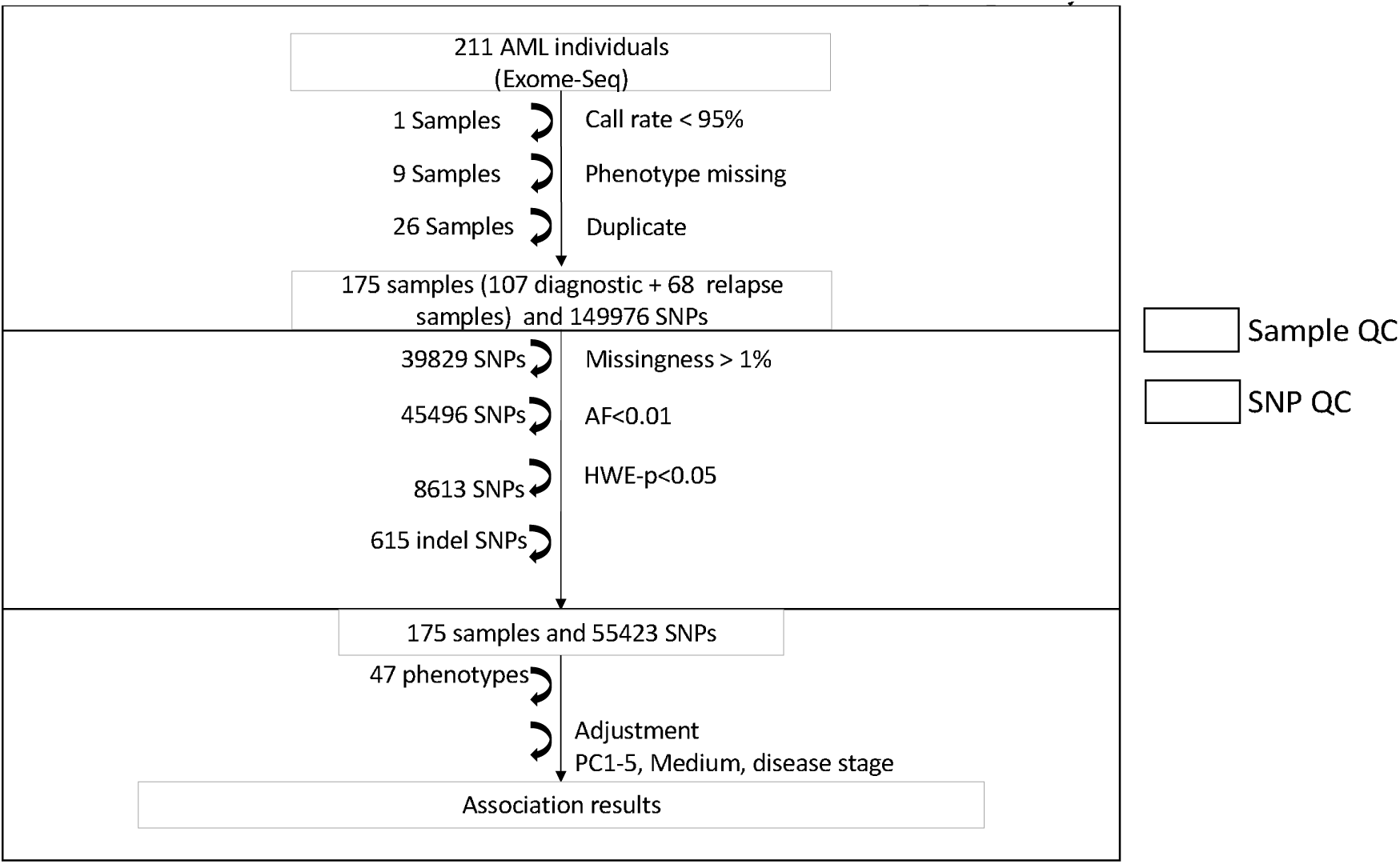
Flow-chart for the SNP and sample quality control.

**Supplementary Figure 2:**
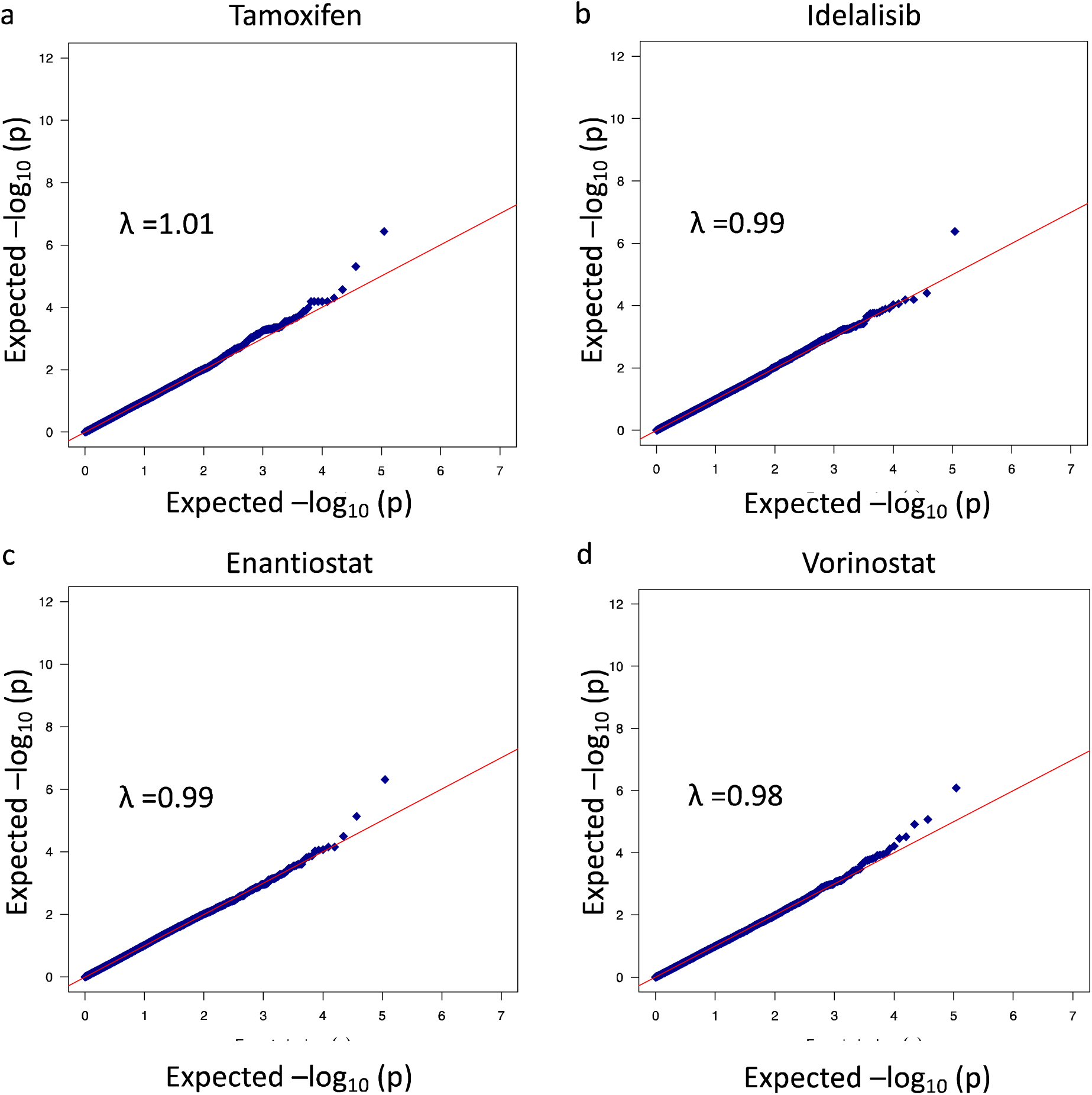
QQ plot for the univariate GWAS associations.

**Supplementary Figure 3:**
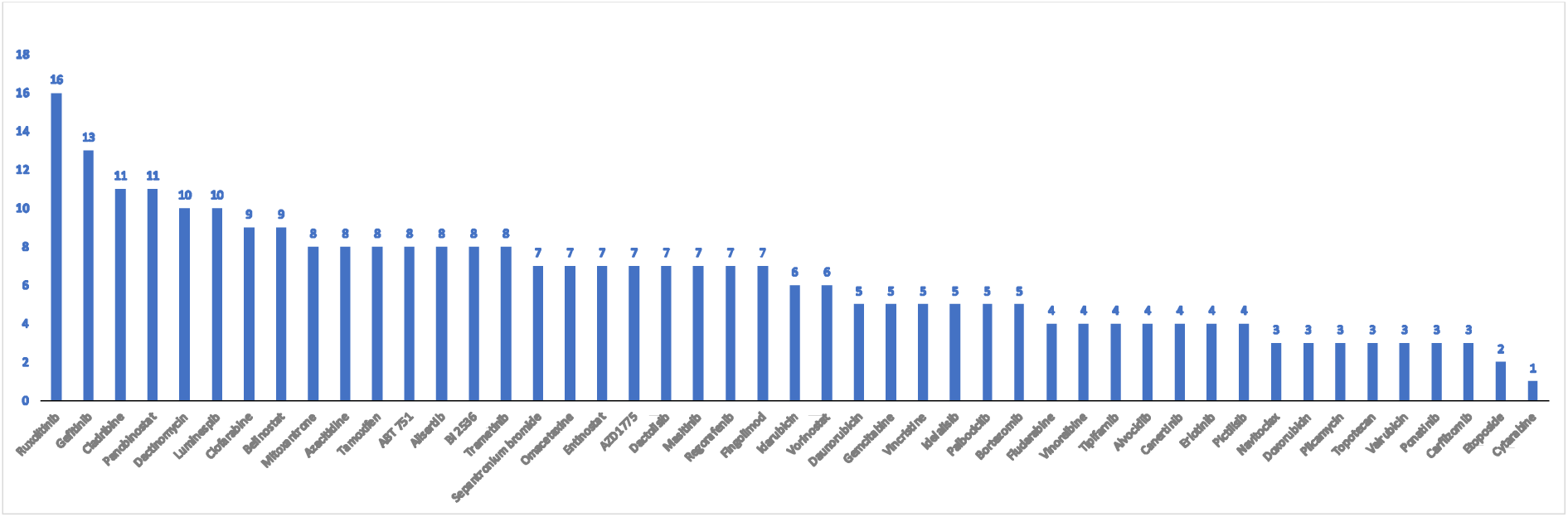
The number of associations observed for each trait at sub exome wide significance level (p<10^-4^).

**Supplementary Figure 4:**
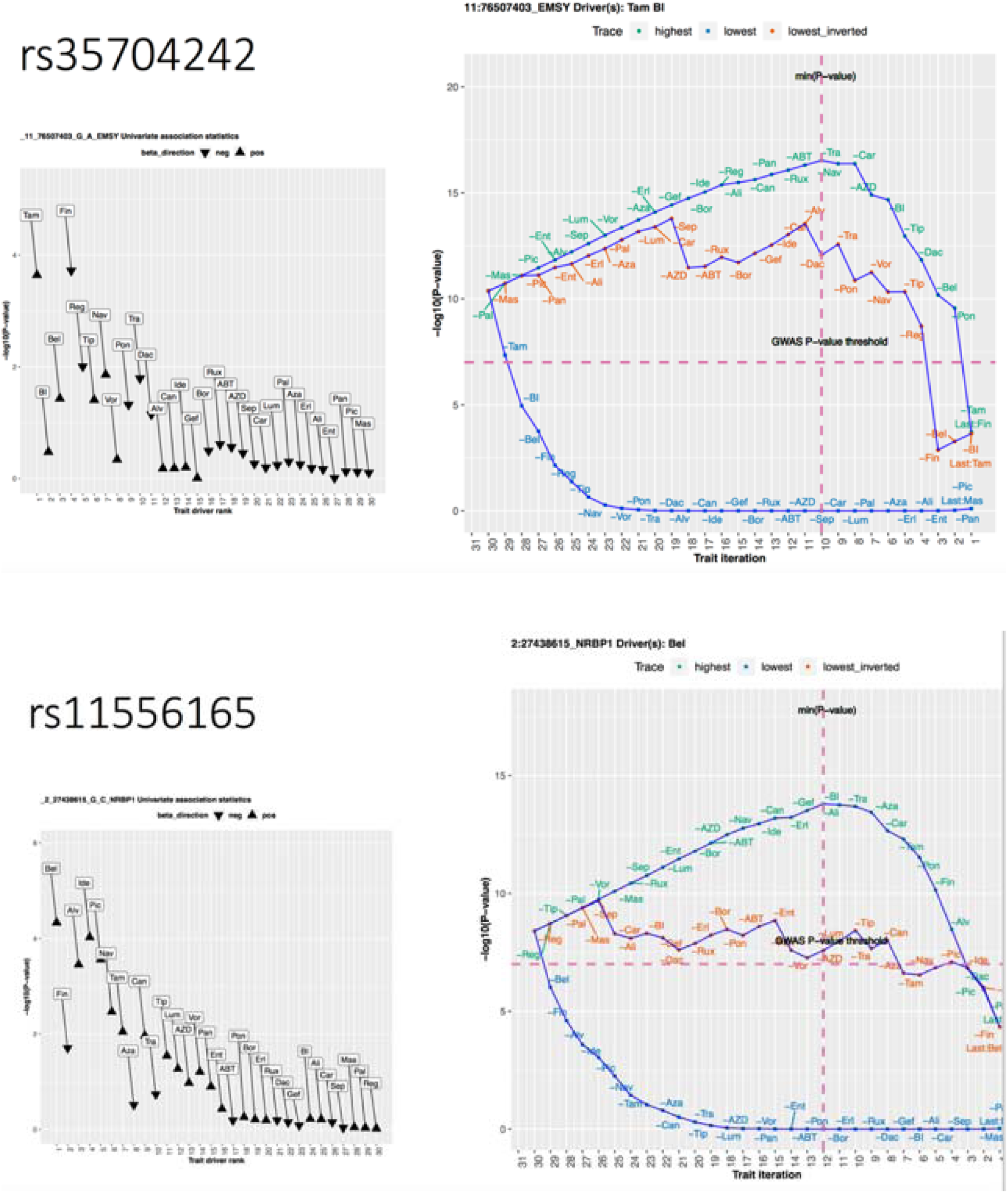
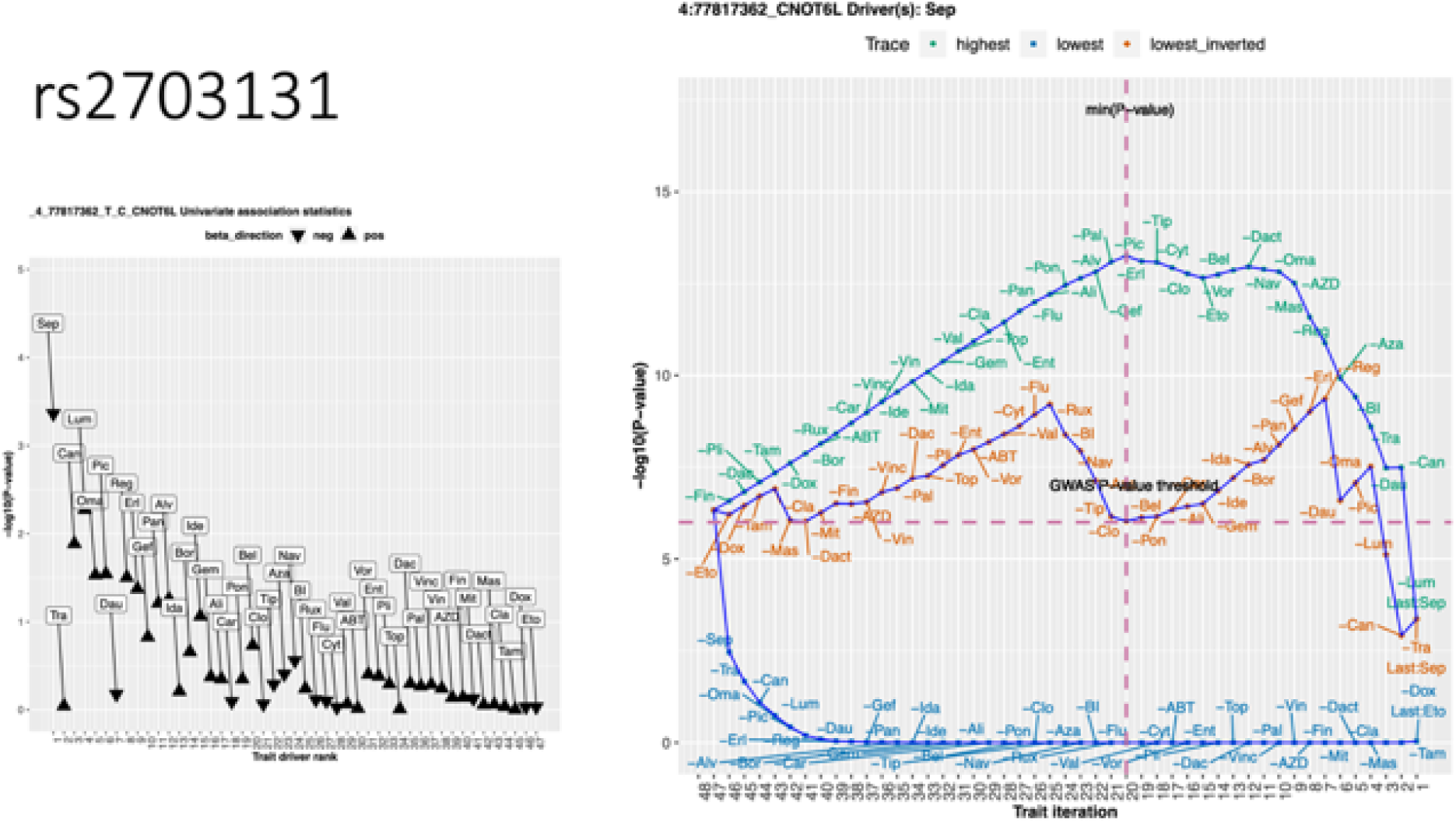
QQ plot for the multivariate analysis.

**Supplementary Figure 5:**
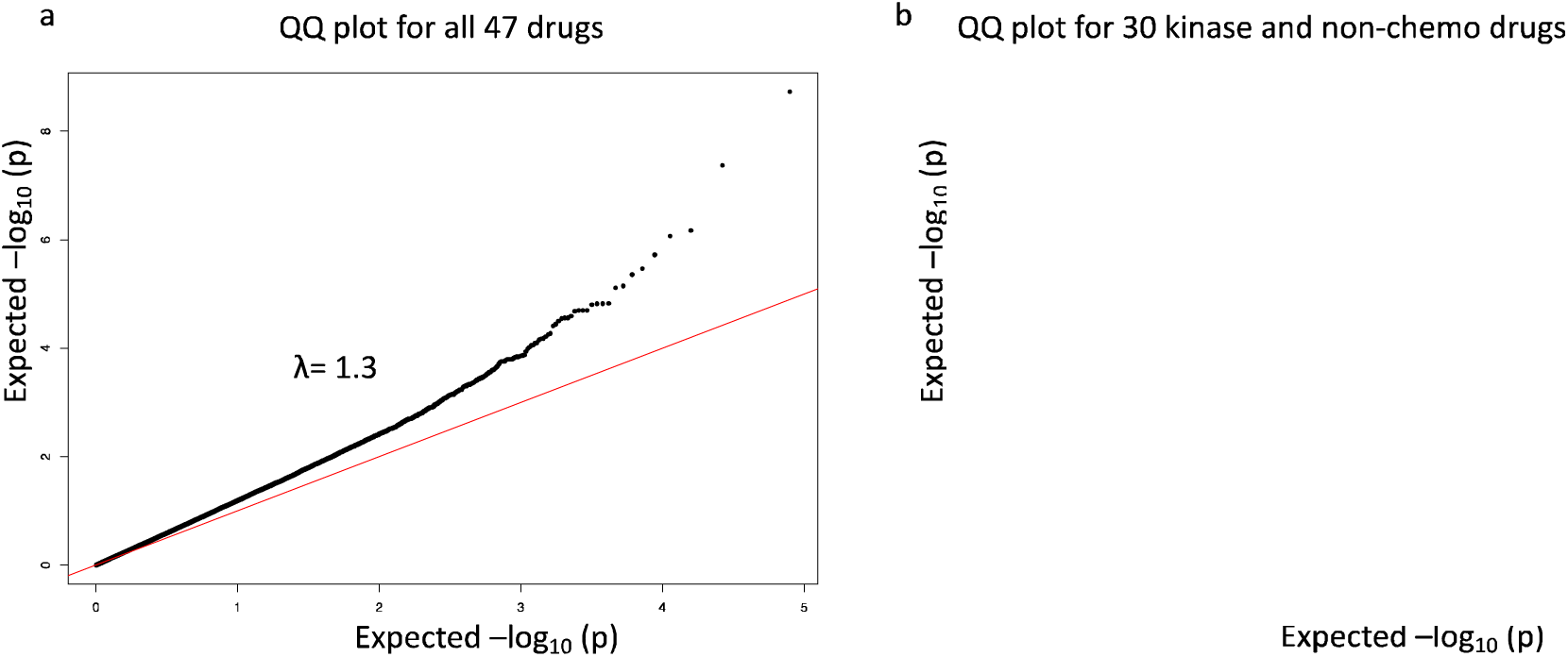
QQ plot for the multivariate analysis.

**Supplementary Table 1:**
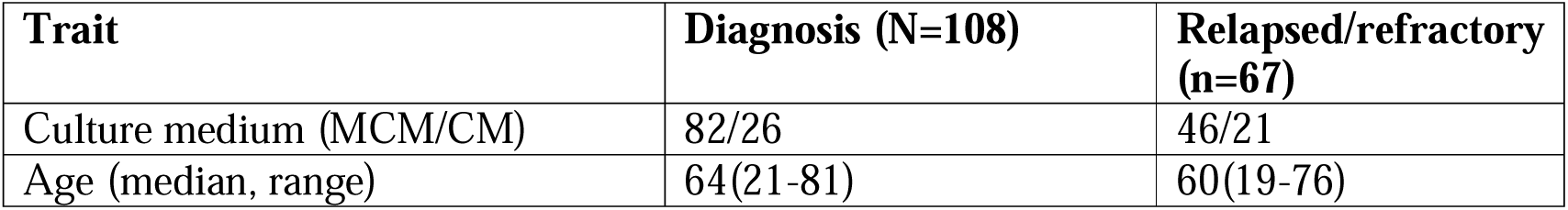
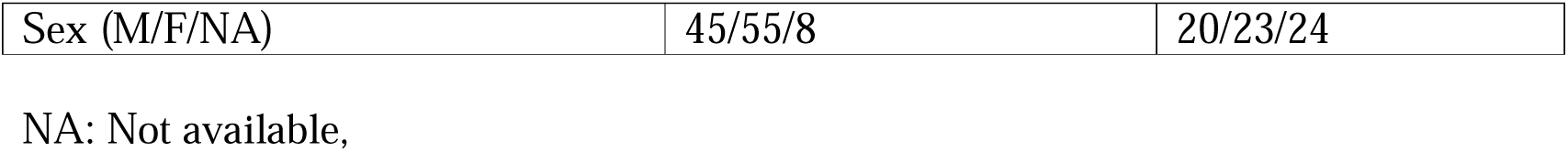
Details of the analyzed drugs and their mechanism of action.

**Supplementary Table 2:**
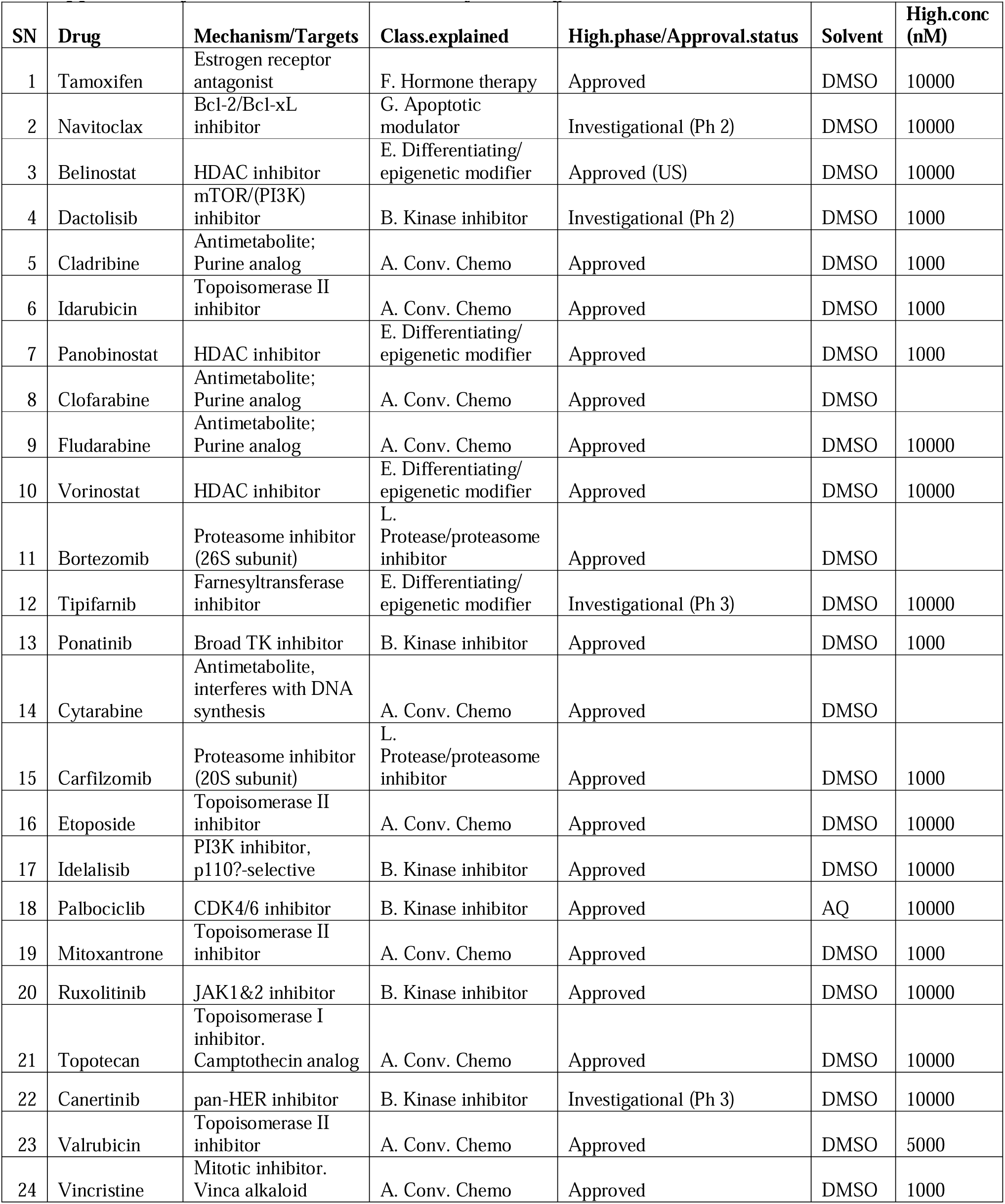
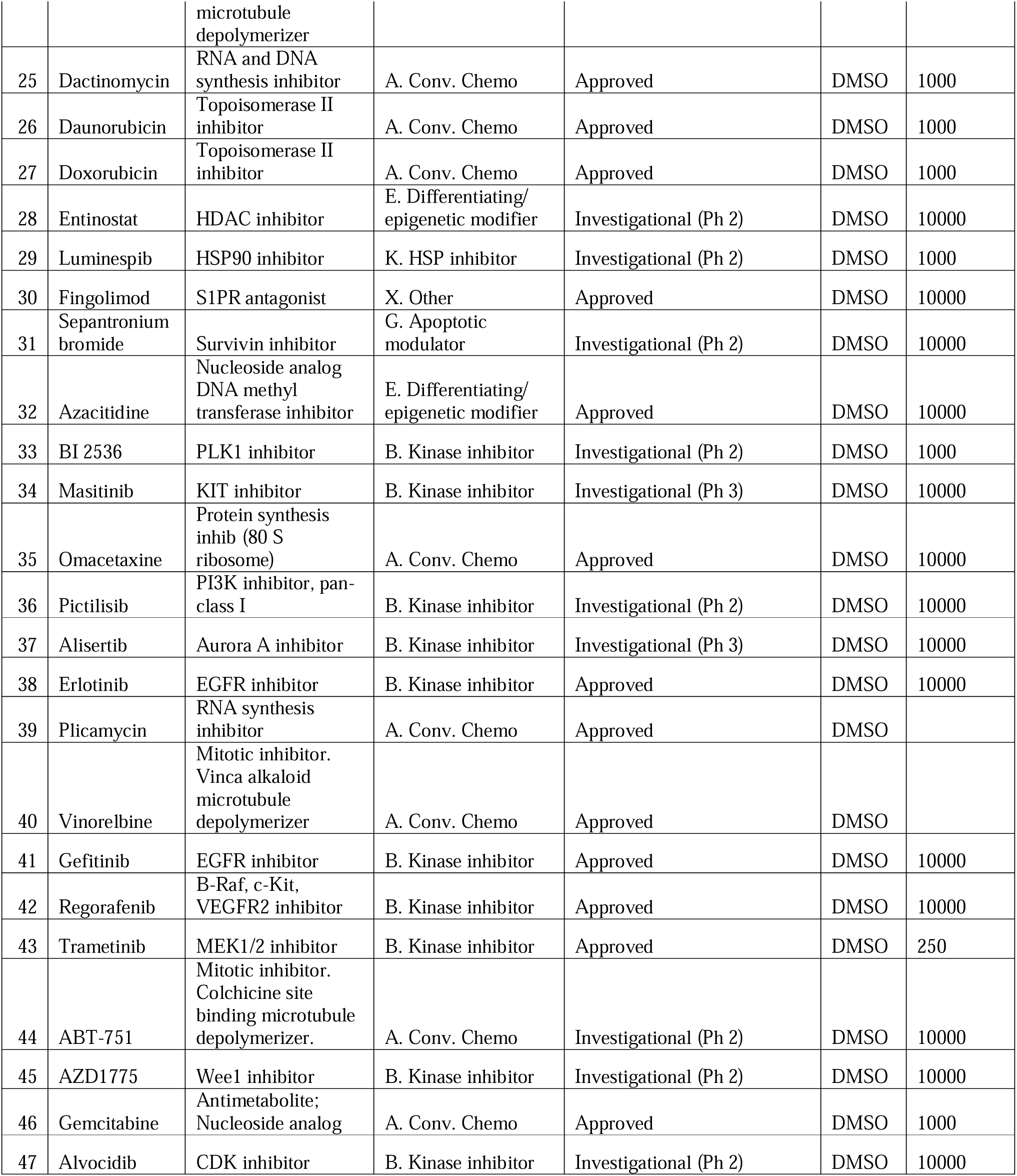
Details of the analyzed drugs and their mechanism of action.

**Supplementary Table 3:**
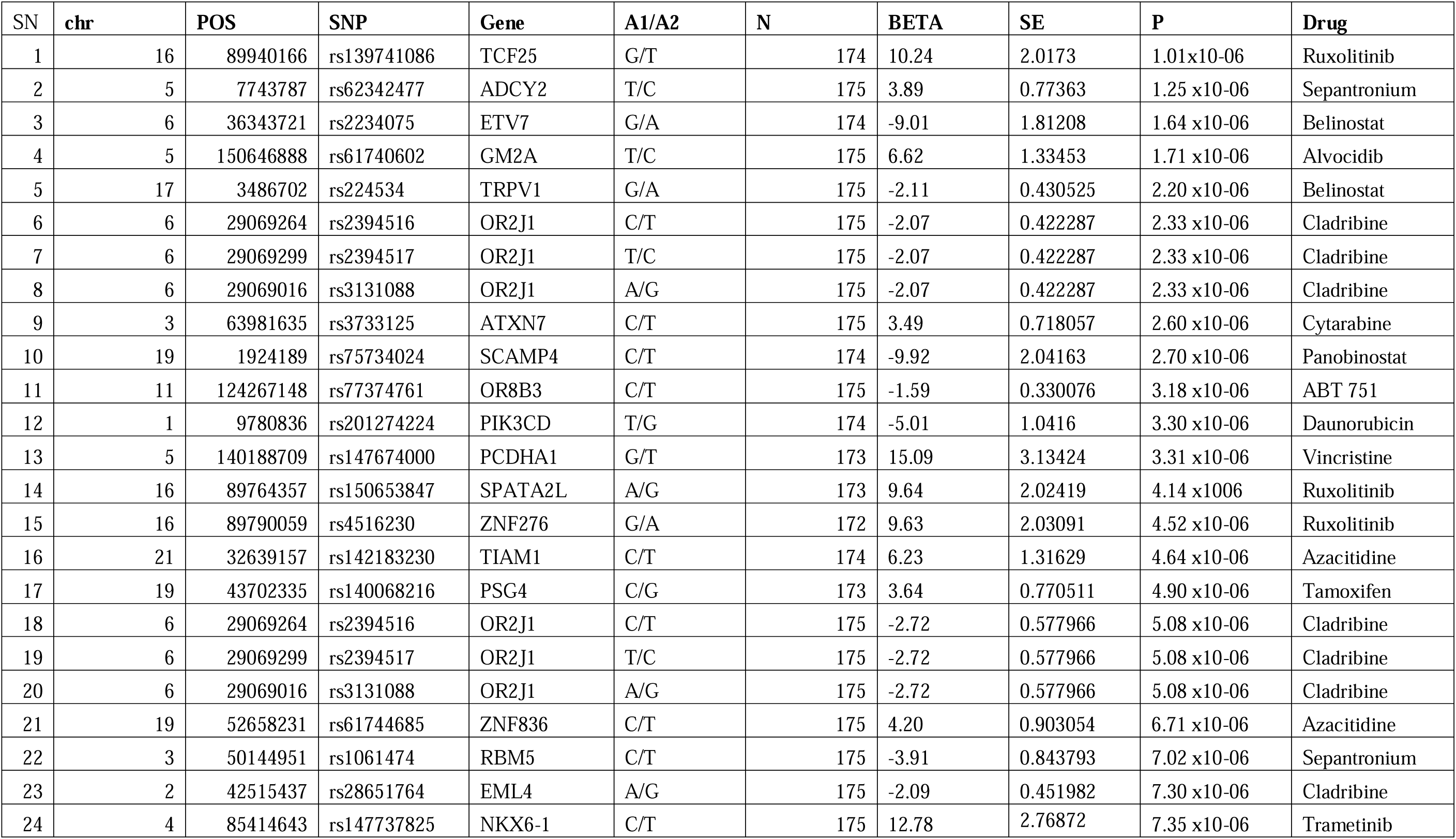
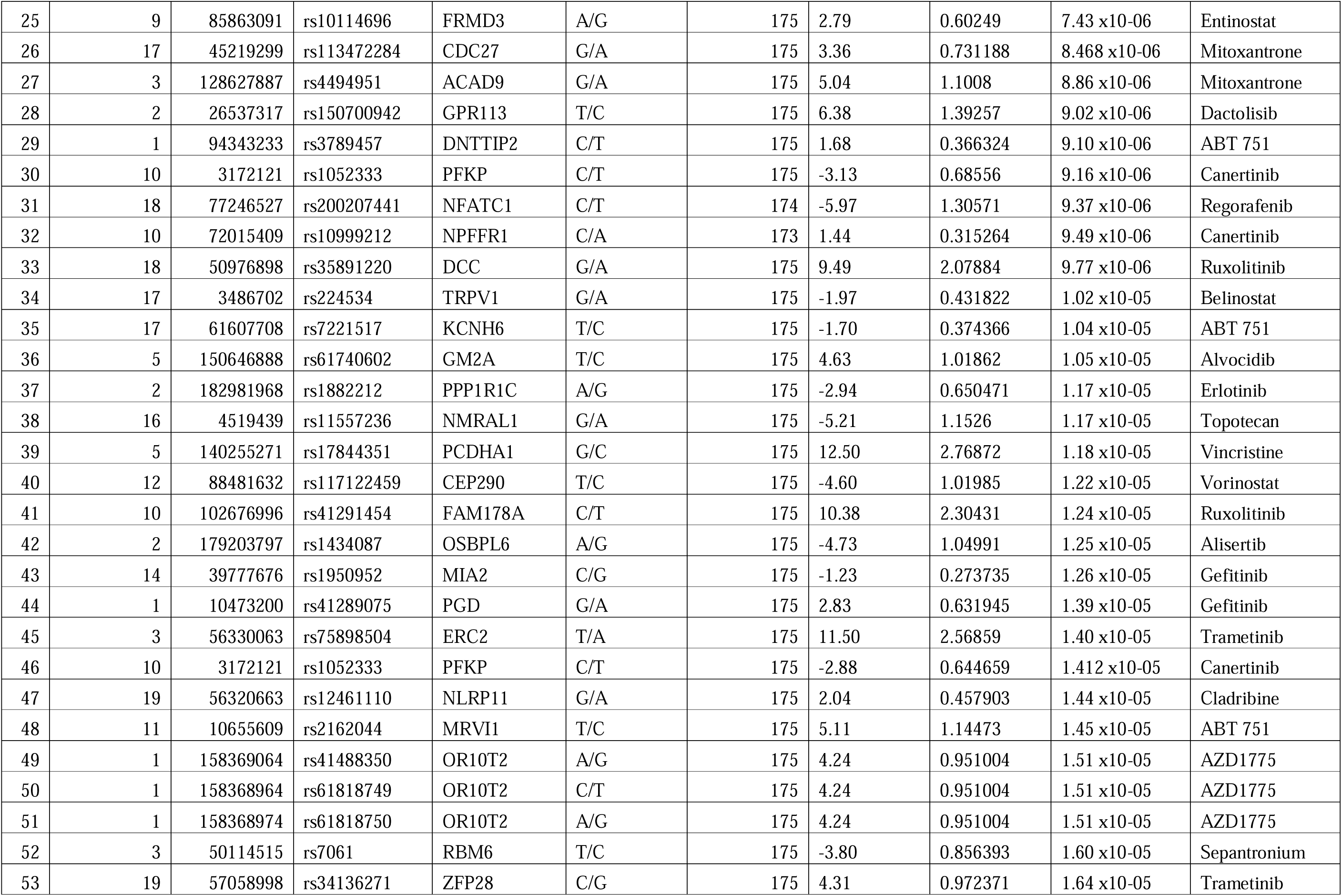
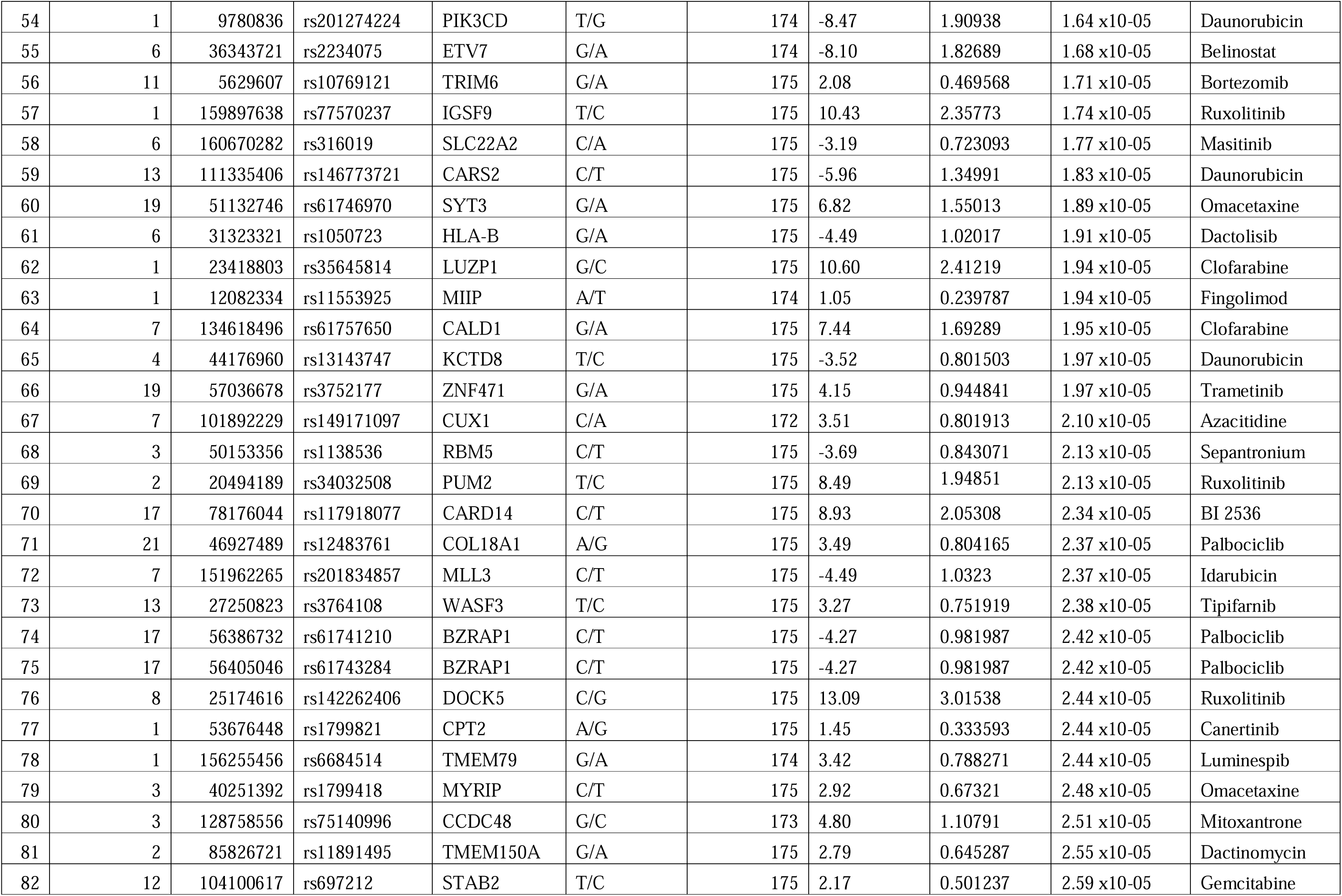
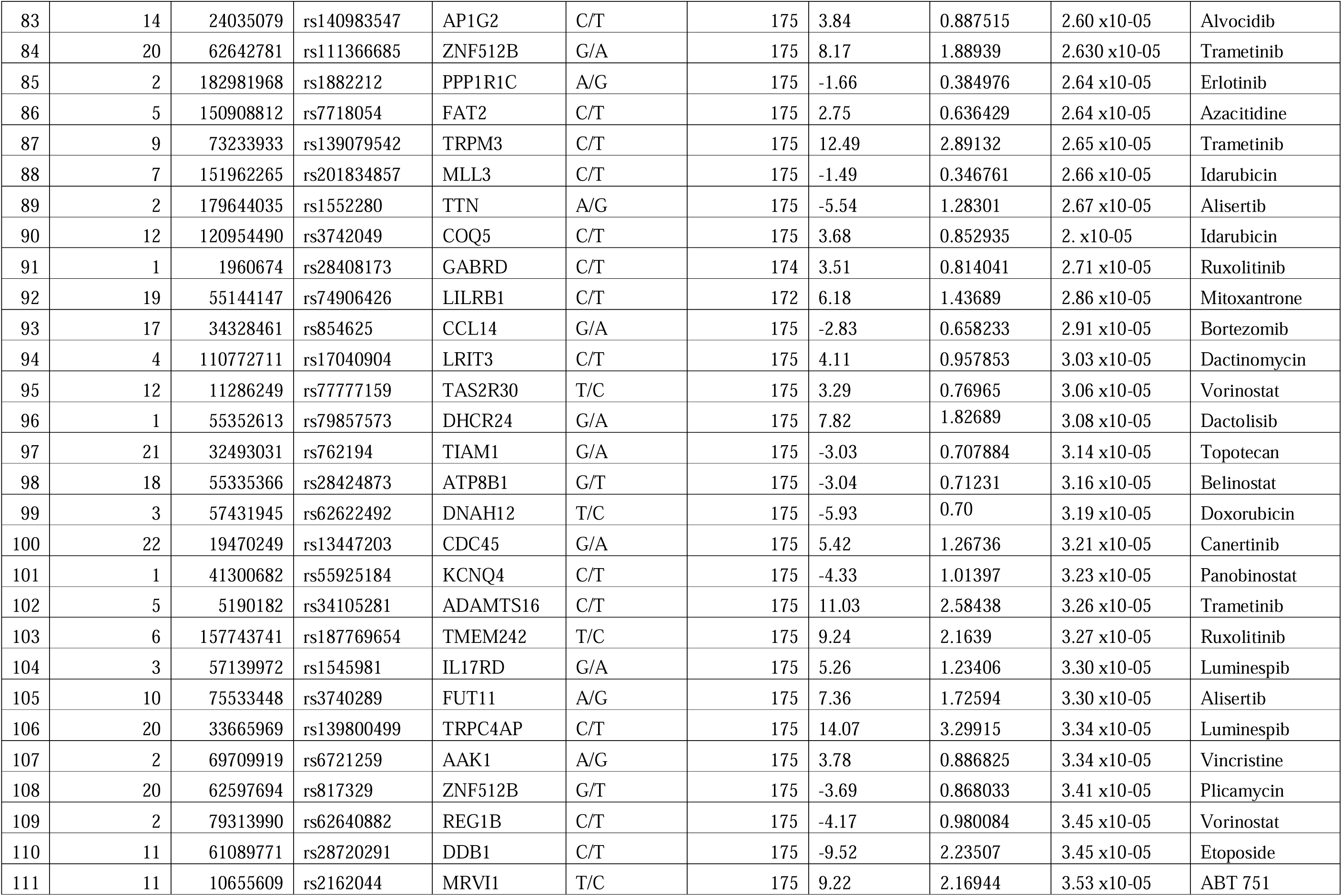
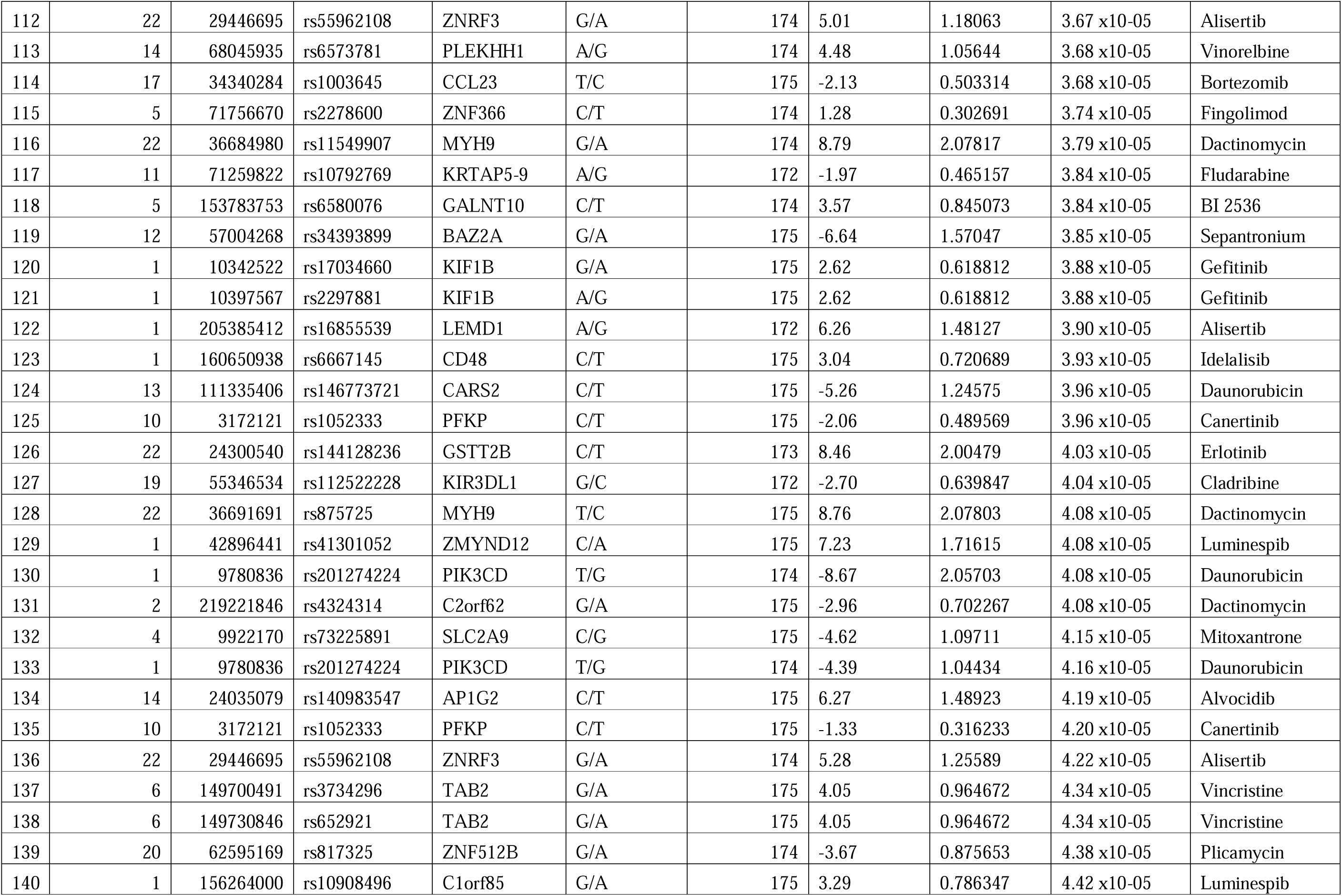
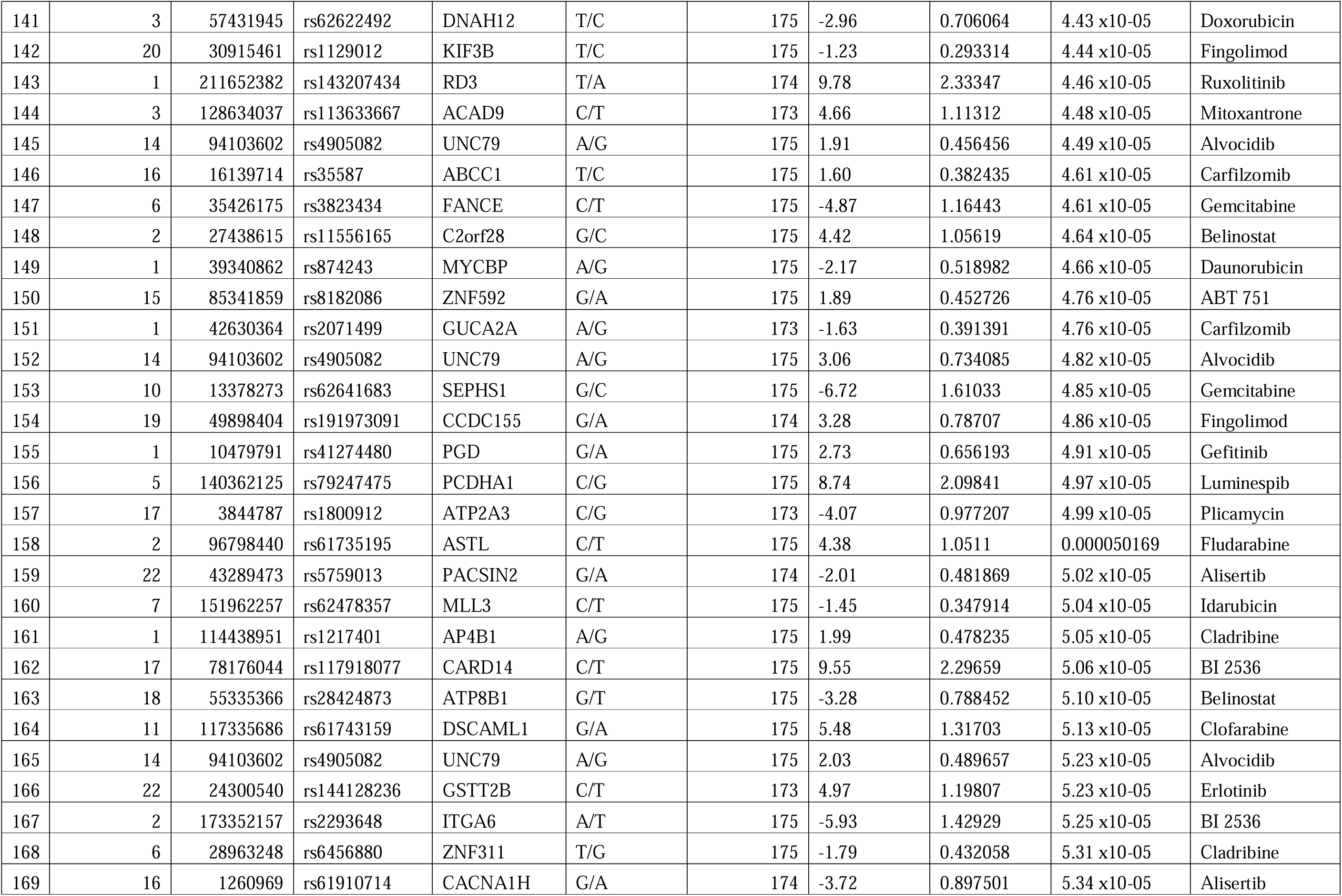
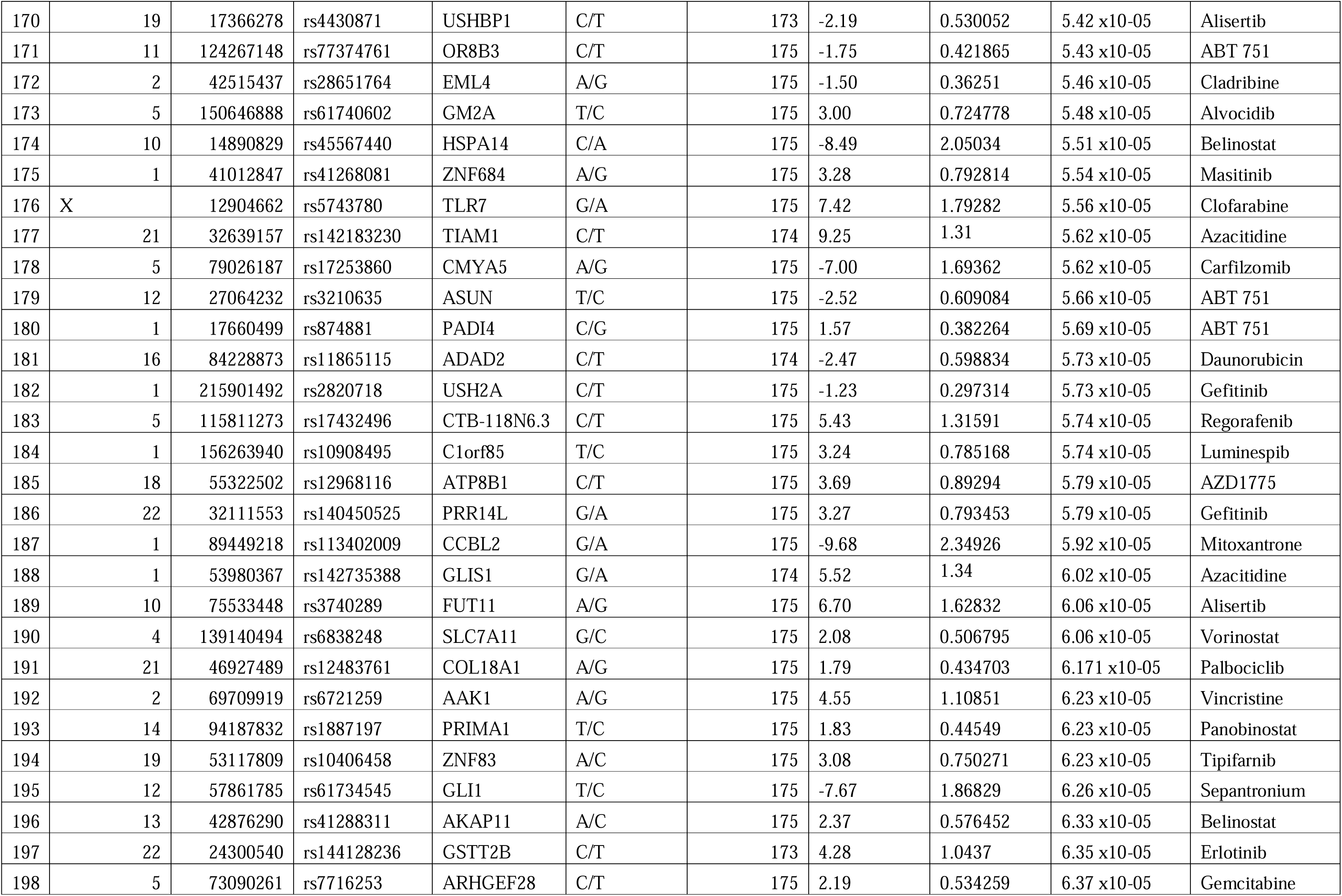
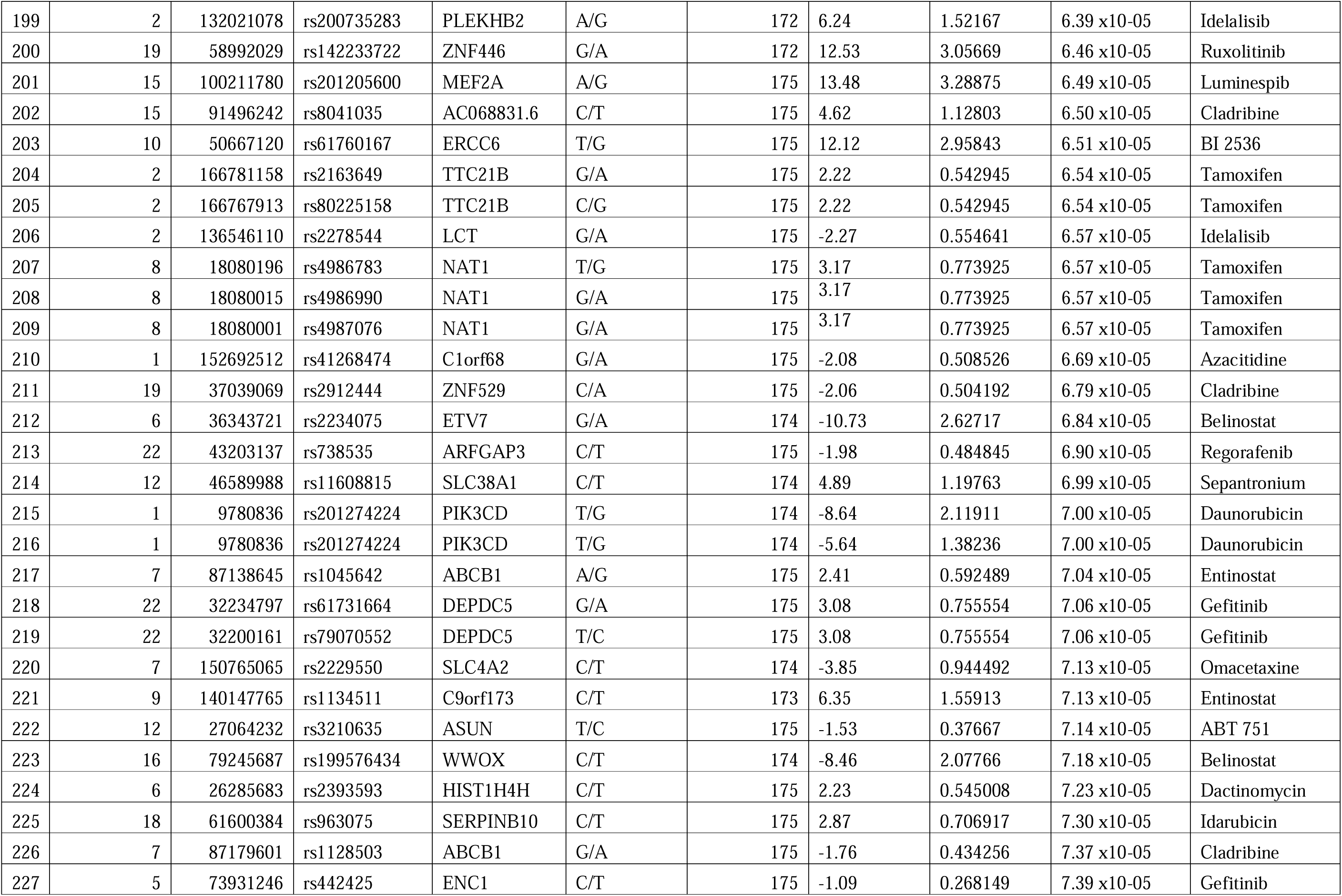
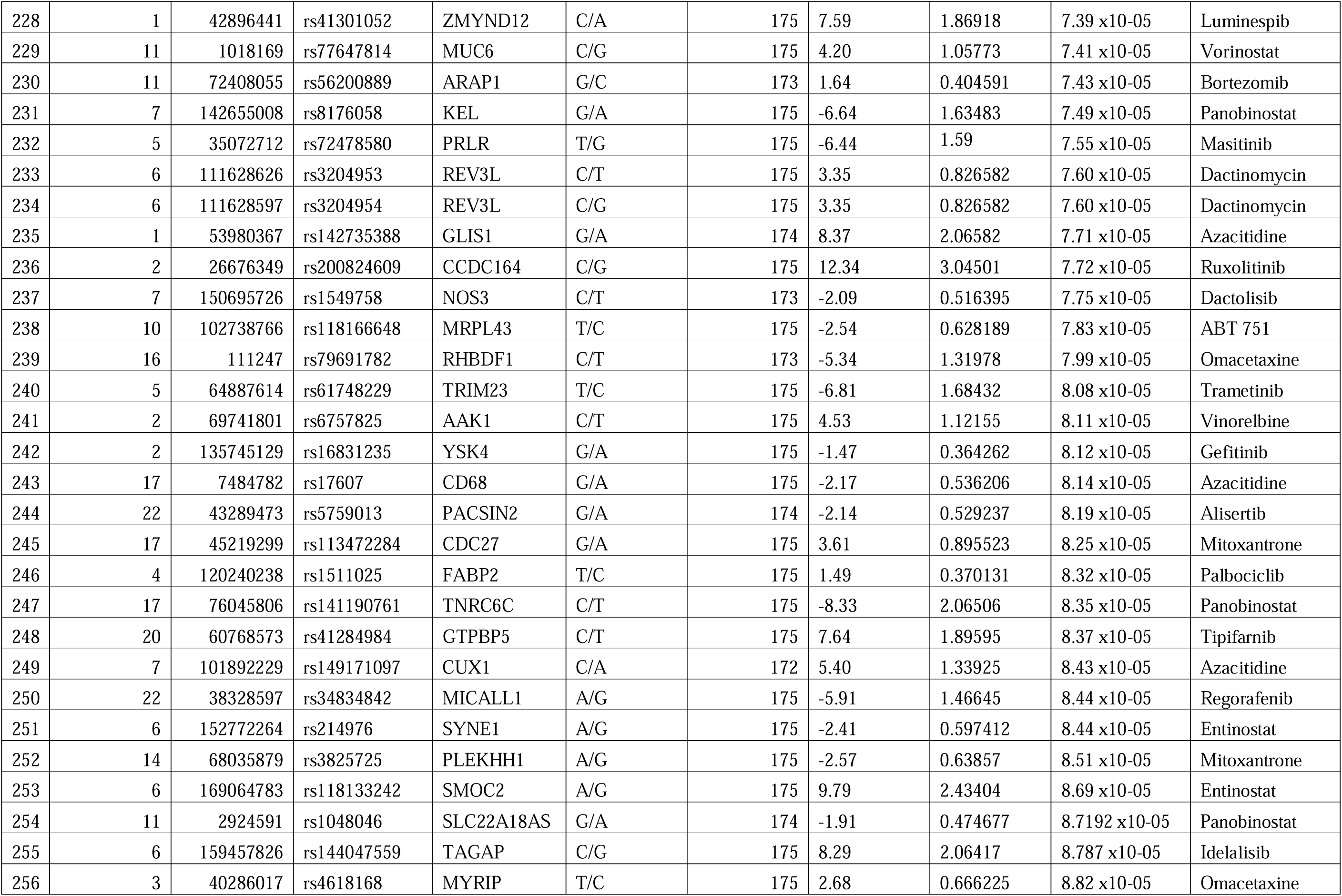
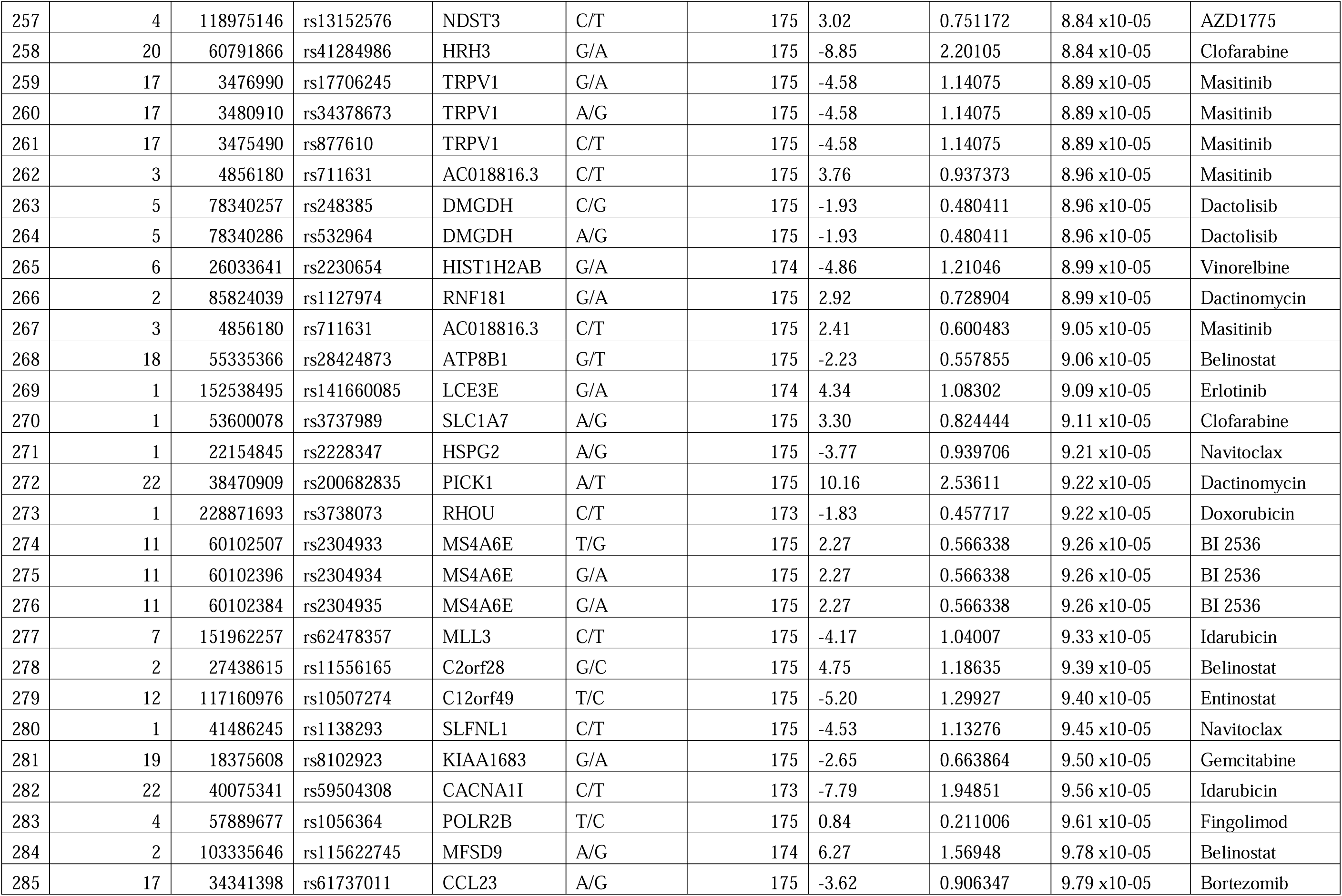
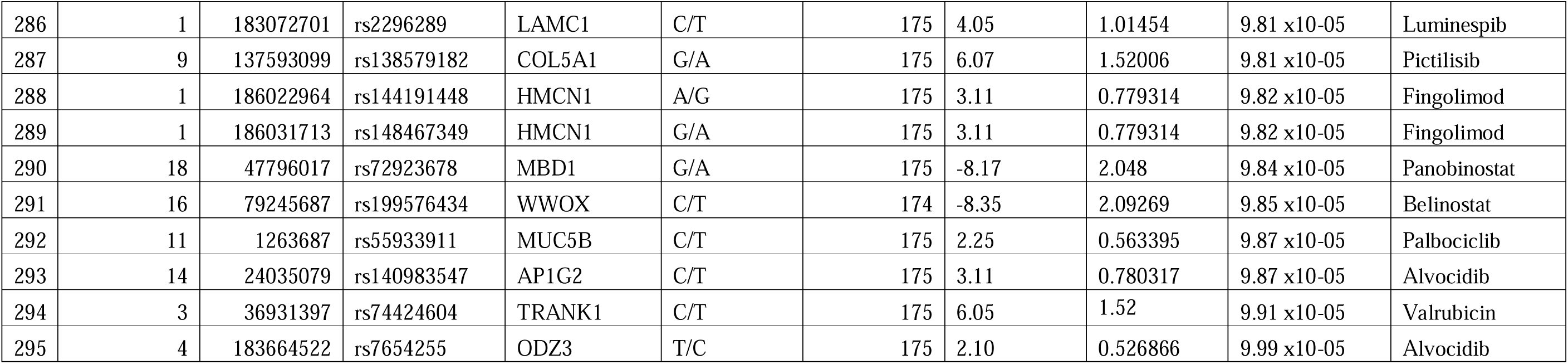
SNPs showing sub-exome wide significance (p<10^-4^) level association with ex-vivo response of drugs in the single variant analysis.

